# A quantitative comparison between human experts and AI at estimating tumor-stroma ratio

**DOI:** 10.1101/2025.08.26.25334451

**Authors:** Ajey Pai, Joren Brunekreef, Eric Marcus, Joyce Sanders, Marcos Da Silva Guimaraes, Renee Menezes, Clara I Sanchez, Hugo M. Horlings, Jonas Teuwen

**Affiliations:** AI for Oncology, The Netherlands Cancer Institute, 1066 CX Amsterdam, The Netherlands; Informatics Institute, University of Amsterdam, Science Park 900, 1098 XH Amsterdam, The Netherlands; Computational Pathology Group, The Netherlands Cancer Institute, 1066 CX Amsterdam, The Netherlands; Biostatistics Centre and Division of Psychosocial Research and Epidemiology, The Netherlands Cancer Institute, 1066 CX Amsterdam, The Netherlands; Department of Pathology, Netherlands Cancer Institute, 1066 CX Amsterdam, The Netherlands; Department of Medical Imaging, Radboud University Medical Center, 6525 GA Nijmegen, The Netherlands

## Abstract

The tumor–stroma ratio (TSR) is an established prognostic biomarker across several cancer types, yet its manual assessment remains labour-intensive and subject to inter-observer variability. An artificial intelligence (AI)-based estimate could offer an efficient, consistent alternative for this task. In this study, quantitative comparisons were made between expert humans and an AI model for TSR estimation. Using two independent, multi-institutional histopathology datasets, an Attention U-Net was benchmarked against experienced pathologists. In a subset of the TCGA-BRCA dataset, the AI model demonstrated comparable trends to human consensus for TSR quantification, achieving an intraclass correlation coefficient (ICC) of 0.69. However, the AI model’s TSR scores are on average 5 percentage points higher compared to human scores on this dataset. The AI model was found to be more consistent at estimating TSR than either of the human counterparts, with a discrepancy ratio (DR) of 0.86. Results on an external dataset obtained from the Netherlands Cancer Institute consisting of cases (n=357) from 35 different Dutch hospitals showed that the AI model’s TSR scores are 7 percentage points lower on average compared to the human rater with an ICC of 0.59. To account for the model’s imperfect segmentation performance, we derived an estimate of the ambiguity in AI-based TSR predictions. The results indicate that, despite this ambiguity, the AI not only follows similar trends but also delivers greater overall scoring consistency than manual TSR assessment by humans.

## Introduction

The tumor-stroma ratio (TSR) reflects the proportion of stromal (non-cancerous supportive) tissue relative to malignant epithelial cells, with higher stromal content often associated with worse outcomes in various solid tumors^1−6^. It became prominent following the growing interest in understanding the role of stromal cells in promoting the growth, proliferation, and survival of cancer cells^7,8^. In breast cancers, TSR is of additional prognostic value for systemic therapy^9^. Currently, TSR is visually quantified in 10% increments of stromal percentage within the most invaded field in whole slide images (WSIs) of cancer resections^10−19^. Given the quantitative nature of the task, an AI-driven approach could add value by ensuring consistent, rapid, and scalable assessments within the most invaded field, which is prone to subjectivity.

It is crucial to note that recent investigations into the performance of AI models designed for similar tasks have uncovered clear evidence that AI models end up capturing non-pathological features relating to the scanner type, staining variations and the hospital where the specimen was prepared^20−22^. As a result, regulatory agencies and clinical bodies remain cautious about endorsing AI-based diagnostics without comprehensive validation across diverse patient cohorts, institutions, and imaging pipelines^23^. In certain instances, AI models are initially developed for quantitative tasks such as segmenting tissue regions or identifying specific cells in the WSIs but end up being employed for measuring biomarkers, which are typically scored subjectively by humans^14,24,25^. Due to the reported inter-observer variability among humans, rigorously comparing AI-derived biomarker estimates with human assessments still demands objective quantification even in the absence of an unequivocal ground truth^26−31^. All these problems render standard segmentation performance metrics like the Dice-Sørensen coefficient (DSC) inadequate^32,33^for evaluating the model’s performance at biomarker estimation.

In this study, we perform a side-by-side evaluation of human and AI-derived tumor-stroma ratio (TSR) estimates to bridge the gap between algorithm development and clinical practice. We quantitatively compare AI-generated and human expert-graded TSR values across two datasets, creating paired, case-by-case records that serve as benchmarks for overall performance and detailed agreement analysis. Importantly, human assessments of TSR inherently involve ambiguity due to the task’s complexity and the lack of an objective ground truth. Similarly, although our AI model produces precise point estimates, segmentation inaccuracies introduce ambiguities in the quantification of TSR that the DSC alone does not capture. By examining both datasets under controlled conditions, we aim not only to assess when humans agree among themselves and with the AI model, but also to characterize the subtleties of disagreement and ambiguity in TSR estimation.

By framing our experiments around paired, expert-annotated and model-predicted TSR values across two independent cohorts, we aim to understand the divergence of AI-derived TSR scores from human estimates in routine practice. Additionally, our experiments attempt to reveal when AI predictions fall outside the acceptable limits of human variability thus making them unreliable for routine application. Solving these problems is essential for translating AI segmentation models into robust, scalable tools that clinicians can trust for prognostic decision-making.

## Results

The Attention U-Net segmentation model we trained achieved a validation Dice-Sørensen coefficient (DSC) of 0.81 on the tumor class and 0.75 on the stroma class. In the third class of all other tissues, the model scores a validation DSC of 0.50. Figure 1 shows an example of the segmentation outputs next to the ground truth segmentation in one of the validation images.

**Figure 1.**
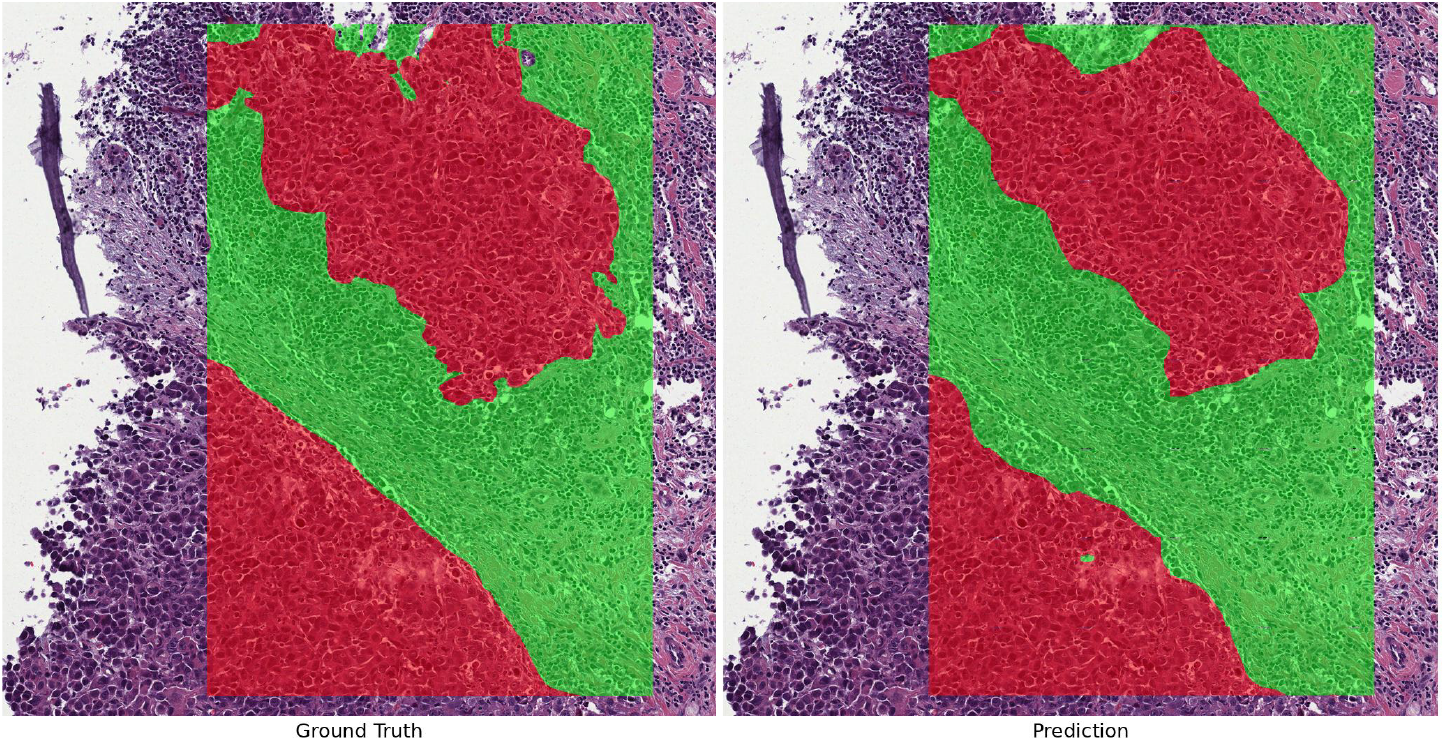
Segmentation output from the Attention U-Net model for tumor and stroma compartments. Left: A region on a whole slide image is shown with polygonal overlay. The regions without an overlay are not used during model training or validation. Right: The corresponding segmentation output from the AI model. Red overlays correspond to tumor regions and green overlays to stroma regions.

In the TCGA-BRCA dataset, Figure 2 shows the trends for the TSR scores provided by the human experts involved in our study (MSG, JS). The Bland-Altman plot (left pane of figure 2) illustrates a systematic difference in the way the observers rate TSR. On average, there is a 14 percentage points difference in their scores. Observer 1 consistently overestimates TSR compared to Observer 2. Additionally, the box plot (right pane of figure 2) suggests that their disagreements are larger between the 30% to 70% range of TSR scores, whereas they agree better in the extreme ranges (0% to 20%, 80% to 100%). The intraclass correlation coefficient (ICC) was 0.66 (95% CI: 0.15−0.84), indicating moderate agreement between the two observers.

**Figure 2.**
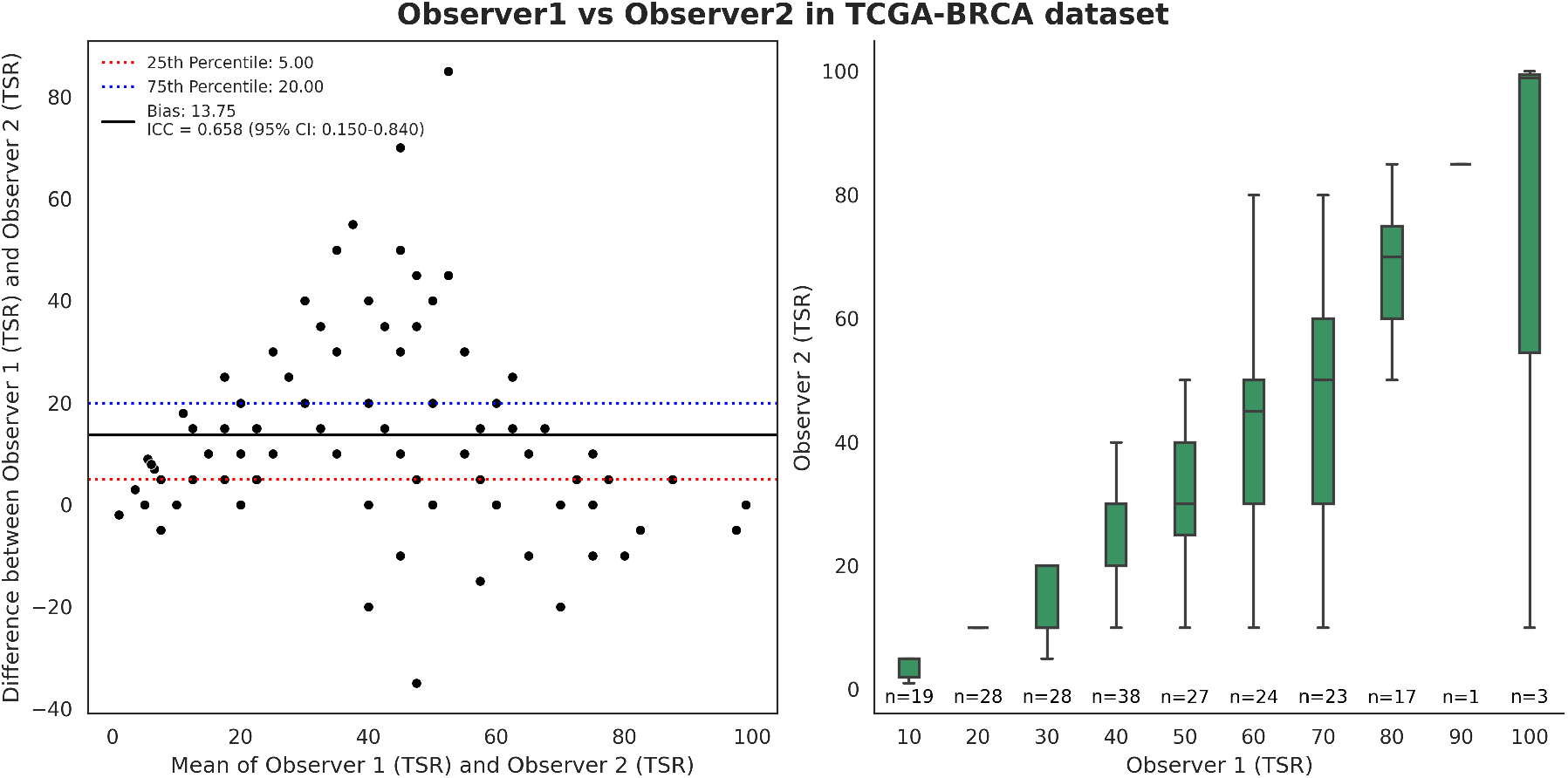
Inter-observer variability in human raters of TSR. The Bland-Altman plot (left) shows a systematic difference of 14 percentage points between the estimates of the two observers. The box plots (right) suggest better agreement between the observers between 0% to 20% and 80% to 100% and a worse agreement in the 30% to 70% range of TSR scores.

In Figure 3, human consensus scores (generated by HMH after examining the individual scores of MSG and JS) for TSR in the TCGA-BRCA dataset are compared with the scores obtained from the AI model. The ICC was 0.69 (95% CI: 0.60−0.76), indicating moderate agreement between human consensus and the AI-derived TSR scores. There is a small bias between the two sets of scores, with the AI-derived estimates being approximately 5 percentage points higher on average.

**Figure 3.**
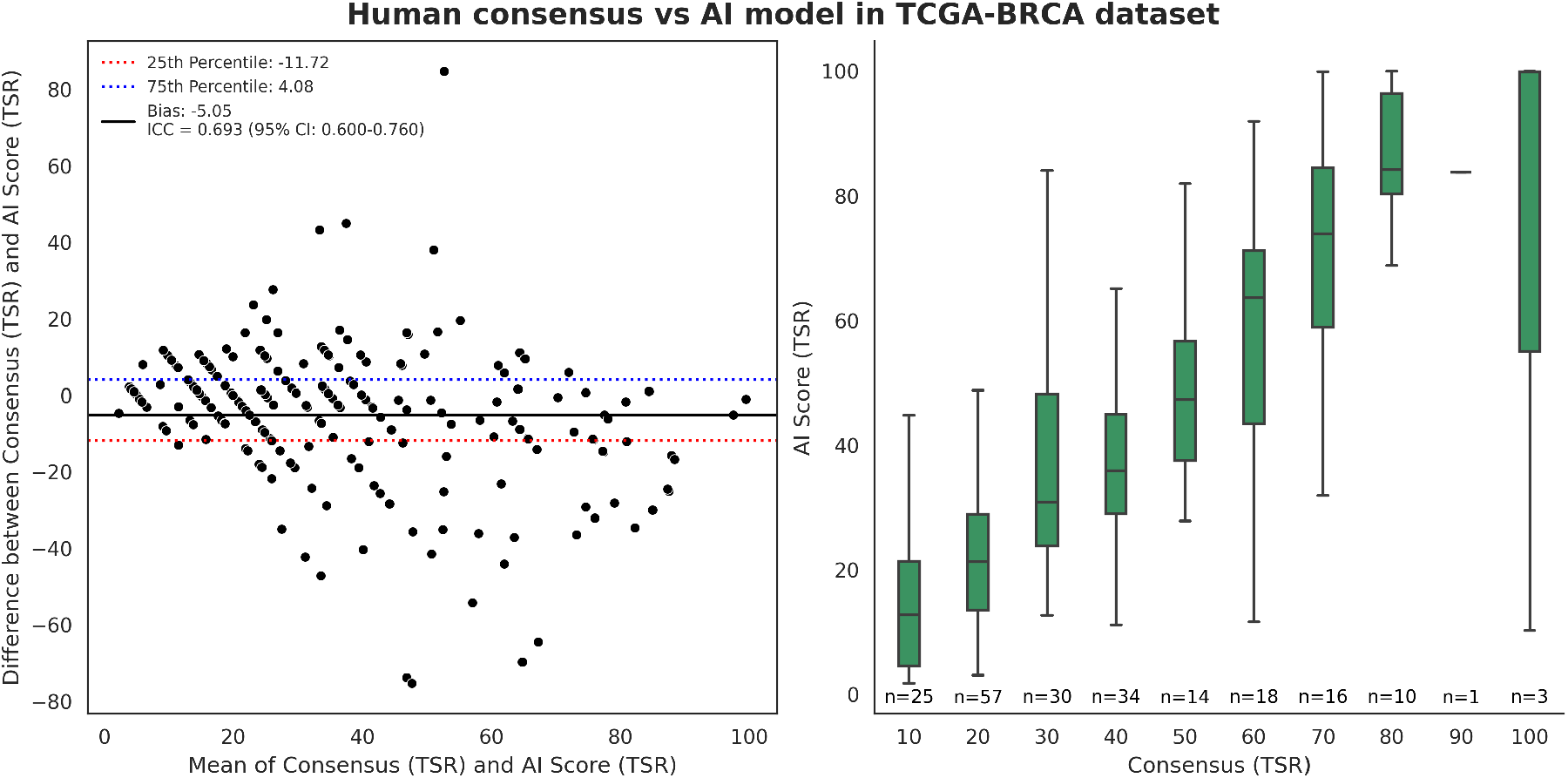
Human consensus vs AI-derived TSR scores

To assess the differences between the AI-derived TSR scores and the individual human observers in the TCGA-BRCA dataset, we employ the discrepancy ratio (DR)^34^. The DR is a metric that measures AI performance against multiple expert humans in the absence of clear ground truth labels for TSR. If the discrepancy ratio is greater than 1, the AI model is less consistent with the average human TSR score than the individual raters. Conversely, if the DR is less than 1, the AI model is more consistent with the average human TSR score than any individual rater. Our AI model has a DR of 0.86, indicating that its TSR estimates align more closely with the human average than the estimates of individual raters.

One of the human experts (MSG) provided TSR scores on a subset of the SmallTNBC dataset (n=357), originating from 35 different Dutch hospitals. These scores were compared against the estimates of the AI model as illustrated in Figure 4. The AI-derived TSR estimates are 7 percentage points lower on average than the human TSR estimates with an ICC of 0.59 (95% CI: 0.49−0.68).

**Figure 4.**
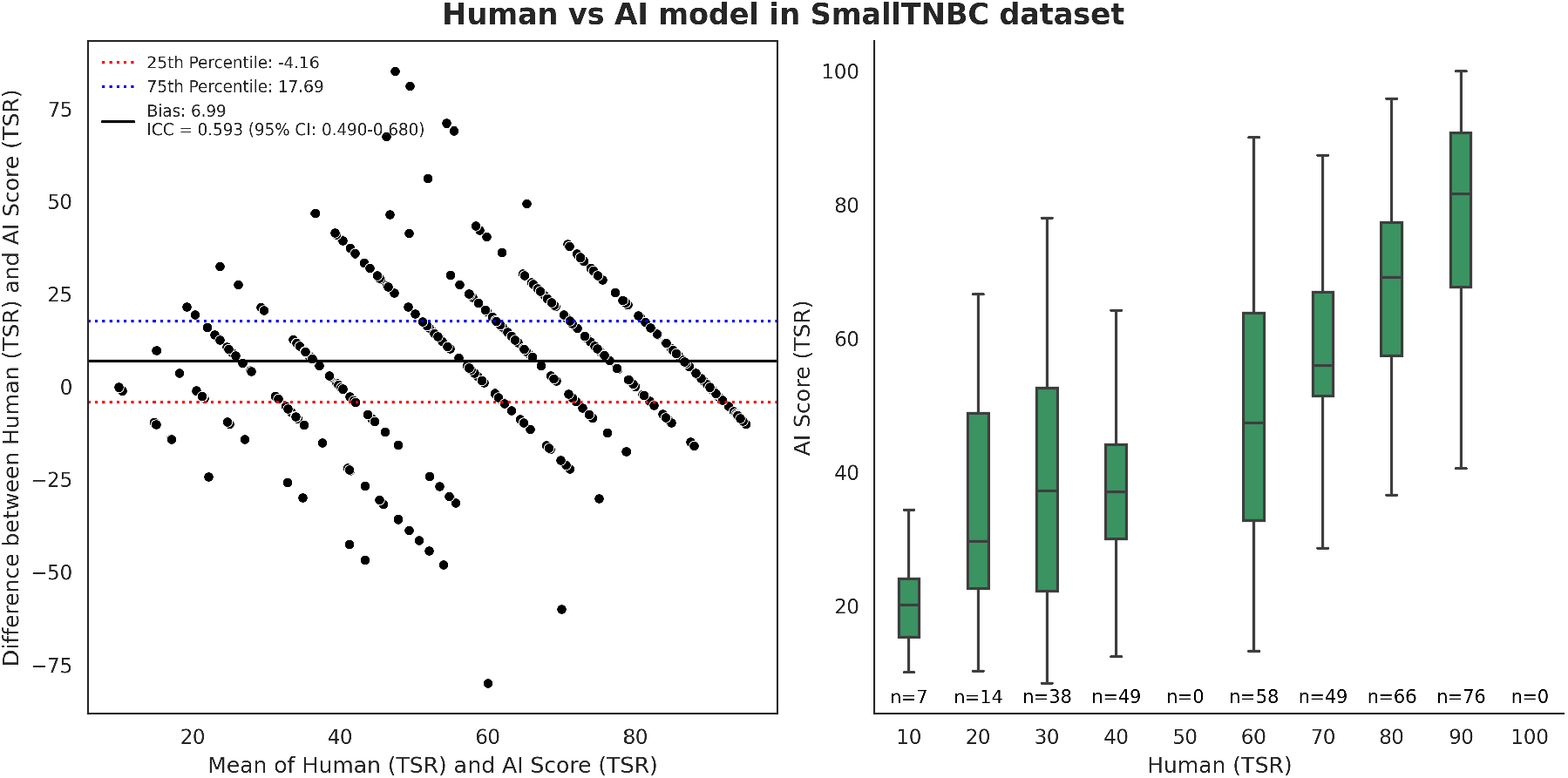
Single human rater vs AI in SmallTNBC dataset

Finally, to ascertain the reliability of the AI model’s TSR predictions, we derived heuristic upper and lower bounds on these TSR values based on the expected segmentation model performance (as measured by the Dice-Sørensen coefficient). This range is what we term the TSR “ambiguity range” (AR). Figure 5 illustrates the behavior of the AI model’s AR across the entire range of TSR scores.

**Figure 5.**
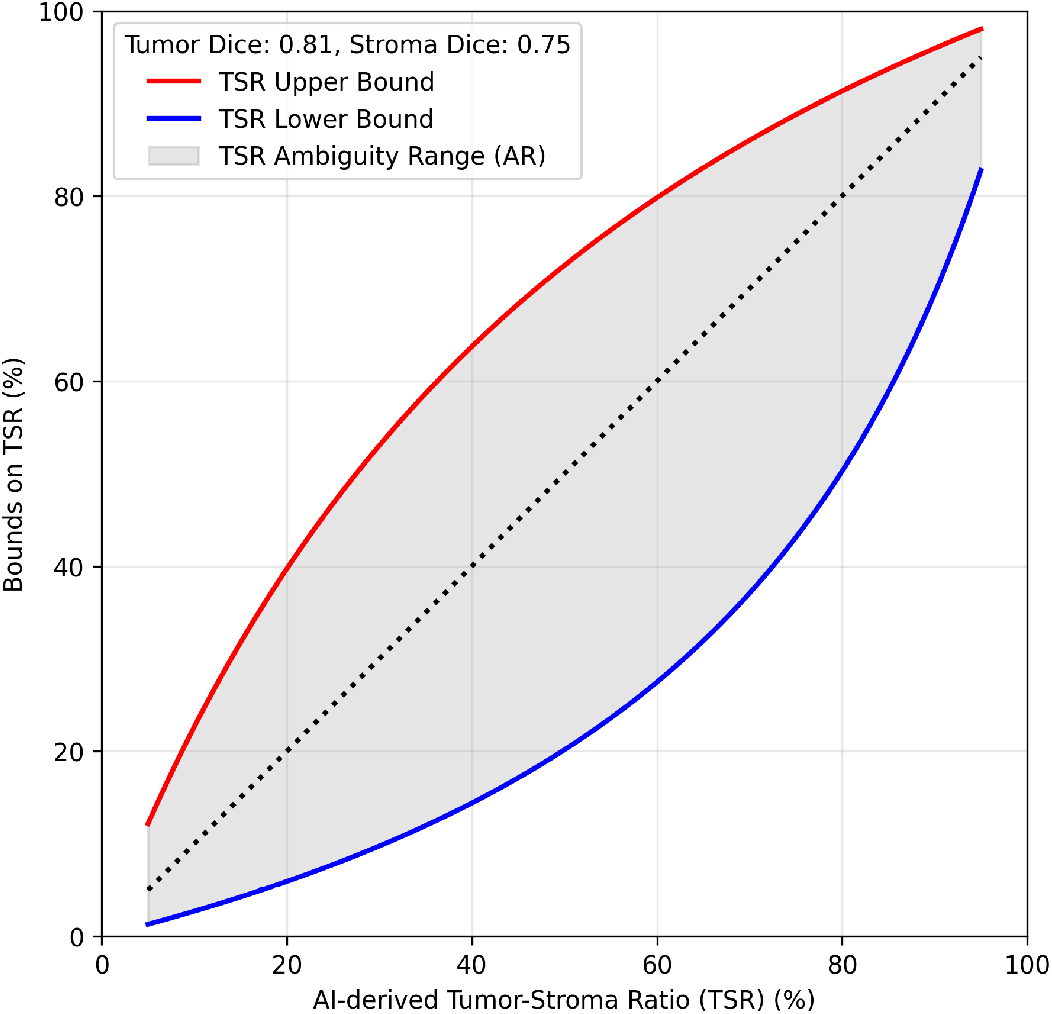
The behavior of the TSR ambiguity range (AR) for different values of AI-derived TSR scores. The ambiguity in quantifying TSR is larger in the intermediate range and tapers off in the extreme ranges.

## Discussion

Our findings indicate that AI-based TSR scoring can surpass the reproducibility of expert pathologists while maintaining only a negligible systematic offset. In the TCGA-BRCA cohort, the model differed from the multi-observer consensus by 5 percentage points on average and in the SmallTNBC subset by 7 percentage points. These biases are smaller than the 14 percentage point systematic gap we observed between the two human observers in TCGA-BRCA, suggesting that variability in TSR scoring stemming from subjective differences among human raters is larger than systematic mistakes made during segmentation by the AI model. The discrepancy ratio of 0.86 supports this observation: each individual pathologist is farther from their colleagues than the model is, implying that the algorithm already behaves like a “third expert” that moderates (rather than amplifies) human disagreements. However, it should be emphasized that the AI performs a pixel-wise segmentation of tumor and stroma components as a first step towards computing the TSR estimates. This is different from the straightforward visual estimation method that humans use to arrive at a TSR score.

Human TSR assessment is a cognitively demanding visual estimation task that relies on implicit heuristics (“eyeballing” cell populations within the most invasive region). In Section 3 of the supplementary material, we illustrate some cases where human estimation of the TSR score is unreliable. The algorithm, in contrast, applies pixel-level segmentation rules consistently across every slide. The resulting algorithmic determinism suppresses the subjective component inherent to human scoring, especially in cases where stromal and tumor compartments intermix and telling them apart is difficult. Our Bland–Altman plots corroborate this: the highest inter-observer disagreement among humans occurs around TSRs between 30–70%, whereas AI-versus-consensus differences remain roughly similar. These observations indicate that algorithmic consistency exceeds that of experts.

Although the intraclass correlation coefficients (ICCs) observed in our study range only from 0.59 to 0.69 — typically interpreted as moderate agreement — this should not be viewed as a shortcoming of the model. Instead, it reflects the inherent ambiguity of TSR estimation, where even human observers demonstrate considerable variability (e.g., ICC = 0.66 between two experts, with a wide 95% CI: 0.15–0.84). The ICC partitions variability into systematic and random components, assuming that disagreement not explained by systematic bias is due to random error. Yet in the TSR estimation task, much of the variability arises from legitimate subjectivity—such as interpretation of poorly demarcated boundaries—rather than noise. This can cause the ICC to underestimate clinically meaningful inter-rater differences or overstate agreement in cases where discrepancies are averaged out.

We also report the discrepancy ratio^34^ of our model’s TSR estimates relative to human observers. The discrepancy ratio was originally proposed to assess AI performance in contexts where experts may disagree on ground truth, and it could prove valuable when commissioning and deploying AI for TSR measurement — for example, in a hospital or diagnostic lab. One pragmatic way to gauge whether the AI is likely to perform reliably is to have at least two pathologists independently score TSR on a small set of WSIs from the target center. The AI-derived TSR scores can then be compared to the human scores via the discrepancy ratio. If the resulting ratio is observed to be below 1, it may be reasonable to consider the model sufficiently aligned with expert opinion for deployment. Our work provides a concrete example how future research in similar contexts can use the discrepancy ratio for assessing model reliability in inference settings.

The TSR ambiguity range (AR) band is a measure that captures how much the AI-derived estimates of TSR could plausibly vary given its measured segmentation accuracy. The magnitude and pattern of AR resemble the variability one observes when multiple expert pathologists independently score TSR, suggesting that the AI’s “ambiguity envelope” (as seen in Figure 5) is a realistic proxy for human disagreement. Potentially, the AR could provide a straightforward commissioning metric: during deployment, end users can predefine an acceptable AR threshold (e.g. 5–10%) and flag any slide whose AI-derived TSR band exceeds that limit for manual review or repeat staining. Beyond triage, AR could also guide model updates by quantifying how much improvement in Dice score is required to shrink the ambiguity to clinically tolerable bounds.

The AI-based TSR prediction approach exhibits several important limitations. First, the underlying segmentation model is never perfect. In Section 3 of the supplementary material, we list several scenarios where the AI model fails at faithfully segmenting the tumor and stroma compartments. The AI model underperforms on out-of-domain instances of data which include rare tissue types, artefacts, and changes in morphology of cells. These problems degrade segmentation quality of the AI model and influence the subsequent TSR estimation step. However, the present study does not aim to correct these failure modes. Instead, we focus exclusively on comparing the AI-derived TSR estimates to those of human experts.

Second, the experiment conducted with the SmallTNBC dataset involves only one rater, as opposed to two raters for the TCGA-BRCA dataset. The lack of a second observer precludes a calculation of the discrepancy ratio. In line with the scope of this work, we did not assess whether the modest AI-human discrepancies would alter downstream survival models.

Third, our TSR ambiguity range (AR) estimate for the AI-based TSR prediction assumes that the model’s Dice-Sørensen coefficient (DSC) on a given sample equals the mean validation DSC — a necessary simplification for deriving the heuristic TSR bounds. By assuming a constant DSC, we ignore errors driven by differences in staining quality, morphological complexity or scanner artifacts. Due to this, it must be noted that the TSR ambiguity range for each sample can be over- or understated.

Overall, our results suggest that segmentation-based AI models offer a rapid alternative for quantifying the TSR by surpassing human consistency and eliminating the need for labor-intensive visual assessment of WSIs.

## Methods

### Datasets

We utilized two datasets — TCGA-BRCA and Histo-AI — to train and evaluate an Attention U-Net^35^ for segmenting tumor and stroma components in H&E-stained whole-slide images (WSIs) of tumor resections. From the public TCGA dataset^36^, we selected a subset of 216 breast cancer (BRCA) WSIs scanned using the Aperio scanner. We obtained ground-truth segmentation masks for various tissue compartments for 151 of these WSIs from the breast cancer segmentation challenge^37^. Furthermore, we selected a subset of 29 P1000-scanned WSIs of breast cancer cases from the Histo-AI pan-cancer dataset collected at the Netherlands Cancer Institute. Ground-truth segmentation masks for the additional 65 slides from the TCGA-BRCA and 29 slides from the Histo-AI were generated by an expert reader and reviewed by a senior pathologist with 25 years of experience. Subsequently, we classified various tissue compartments as either tumor or stroma according to previously established guidelines^18^. Data annotation details are provided in the supplementary Section 1.1.

To evaluate human and AI performance in TSR quantification on an external dataset, we used the SmallTNBC cohort, which comprises WSIs from 2,109 early-stage triple-negative breast cancer (TNBC) patients treated across 35 hospitals in the Netherlands. From this cohort, we acquired TSR scores for a subset of 357 P1000-scanned WSIs. One of the two pathologists who scored the TCGA-BRCA dataset also assessed the SmallTNBC slides, ensuring continuity in expert judgment.

### Ethics

The institutional review board at the NKI-AVL approved the retrospective use of Histo-AI dataset (IRB number: IRBdm20-140) and the SmallTNBC dataset (IRB number: IRBdm20-231).

### Segmentation model training

We trained the Attention U-Net segmentation model using 166 WSIs from 156 patients (144 Aperio from TCGA-BRCA and 22 P1000 from Histo-AI) and validated it on 79 WSIs from 76 patients (72 Aperio and 7 P1000). Out of the 216 WSIs from the TCGA-BRCA dataset, we used 209 WSIs to compare the model’s tumor–stroma ratio (TSR) estimates with TSR estimates from two pathologists (MSG has 12 years of experience and JS has 23 years of experience). The sub-regions used for TSR scoring were distinct from the sub-regions used for training and validation of the segmentation model, ensuring that data leakage would not bias our results. We evaluate the segmentation model’s performance on the validation set with the Dice-Sørensen coefficient (DSC) for both the tumor and stroma compartments. Further details on model training and evaluation can be found in supplementary Section 1.

### Estimation of TSR

Three pathologists were involved in estimating TSR scores in this study. One of the pathologists (HMH) acted as adjudicator who initially determined the stroma-rich most invasive region (MIR) for each WSI in the TCGA-BRCA dataset following prior guidelines^18^. Subsequently, all the MIRs in the WSIs were scored for TSR by two other pathologists (MSG, JS) independently. Finally, the adjudicator (HMH) proposed consensus scores by looking at the TSR estimates of the other two experts. Slides were scored through the online scoring platform Slidescore^38^. We performed inter-observer agreement analysis on the TSR estimates thus obtained from expert humans.

Once the segmentation maps from the AI model were obtained, the TSR scores within the MIRs were computed as a post-processing step by counting the number of pixels classified as tumor or stroma categories. We furthermore compared human consensus with the AI-derived TSR scores on the TCGA-BRCA dataset. The tumor-stroma ratio is defined as

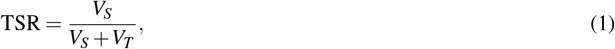

where *V_S_* and *V_T_* are the numbers of pixels classified as stroma and tumor, respectively, by the AI model in the MIRs. We summarize the TSR scoring workflow for both the AI model and the human rater in Figure 6.

**Figure 6.**
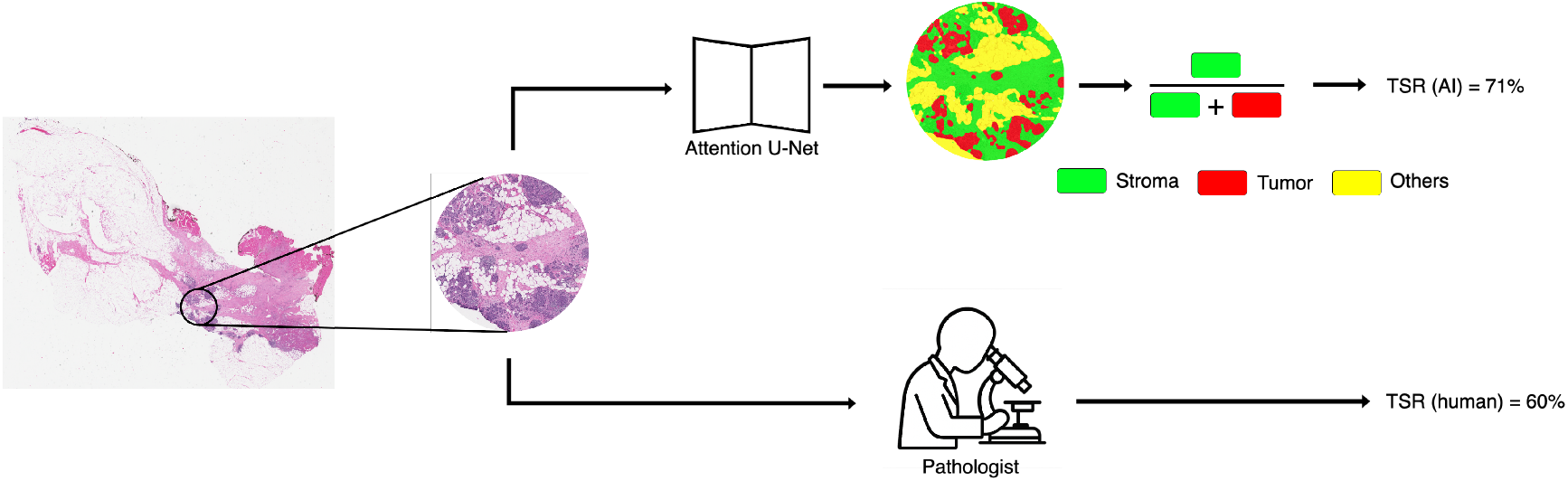
AI and human scoring procedures. In the AI workflow, a segmentation model first segments various tissue compartments in the most invasive region (MIR). The pixel counts for the tumor and stroma compartments are aggregated into a tumor-stroma ratio (TSR). The human pathologist produces a TSR estimate directly from a visual inspection of the MIR.

In order to investigate human and AI performance on an external cohort, we first followed the aforementioned procedure to extract AI-derived TSR scores on the SmallTNBC dataset. MSG provided TSR scores in the MIRs on this dataset and these scores were subsequently used for comparing human and AI performance.

### Comparing humans and AI in the absence of ground truth for TSR

In real-world settings where AI models are deployed for inference, it is important to determine the reliability of the AI model against expert humans. However, the subjective nature of TSR scores generated by humans makes it difficult to perform an objective comparison to the AI-derived scores. In such scenarios, we can rely on the discrepancy ratio (DR), which is a metric designed to compare expert humans and AI models in the absence of unambiguous ground-truth labels^34^. The DR is defined as:

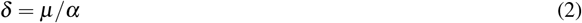

Where (*α*)?is the average difference among all human observers, (*µ*)?is the average difference between the AI model and each of the human observers. The DR represents the ratio between the average disagreement of the AI model with each human observer and the disagreement among human observers. A DR greater than 1 indicates that the model is more inconsistent with its estimates compared to the average human observer. On the other hand, a DR of less than 1 indicates higher consistency of the AI model compared the average human observer.

On 209 WSIs from the TCGA-BRCA cohort, we conduct a reader study where two humans (MSG, JS) independently estimate TSR in MIRs marked out by a third expert (HMH) who also acts as the adjudicator. To measure the DR, the differences between individual human assessments and the differences in AI-derived scores and individual human ratings for the TSR are measured using the absolute distance.

### Heuristic bounds on AI-derived TSR predictions

The disagreements in the TSR estimates of human observers are caused by differences in how each observer analyzes the samples. The correctness of a single observer’s assessment cannot be established due to the lack of an objective ground truth. Therefore, the human TSR estimates are subject to a certain inter-observer variability, which is apparent in the spread of the TSR predictions obtained for each individual sample.

Our AI-derived TSR estimates are — on a first impression — point predictions, without any notion of uncertainty or margin of error. However, we base these TSR estimates on a mask that was generated by an AI segmentation model. Such segmentation models are generally imperfect, meaning that they assign incorrect labels to some pixels from the input image. The Dice-Sørensen coefficient (DSC) quantifies how frequent these pixel misclassifications are in one particular image. The average DSC of the segmentation model on each of the images in the validation set therefore provides an estimate of the number of misclassified pixels in unseen tissue regions. Recognizing that the TSR is computed as a ratio of pixel counts (as seen in equation 1), we can formulate a heuristic notion of ambiguity on an AI-derived TSR estimate by taking into account the expected number of misclassified pixels. This leads to the following upper and lower “bounds” for the AI TSR estimates (we refer to supplementary Section 2 for a derivation):

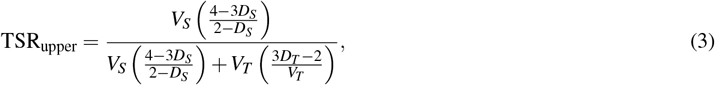

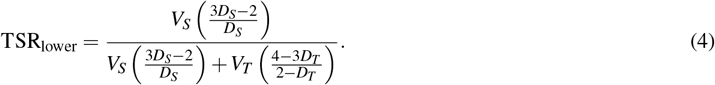

where *V_S,T_* are the numbers of pixels and *D_S,T_* the average validation DSC coefficients for the stroma and tumor compartments, respectively. We refer to the range of these bounds as the TSR ambiguity range (AR) of the AI model:

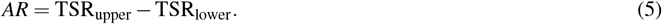

We point out that these bounds do not correspond to confidence intervals in the classical, statistical sense and they should not be interpreted as such. They are rather heuristic bounds that provide a range of TSR estimates for a given tissue region that are consistent with the model’s segmentation output, assuming that the model’s performance on this tissue region is similar to its performance on the validation images.

### Statistical analysis

ICC estimates and their 95% confidence intervals were calculated using pingouin statistical package for python (version 0.5.5) based on a mean-rating (k = 2), absolute-agreement, 2-way random-effects model^39^. Prior guidelines for interpreting the ICC have been followed in this work^40^.

## Data Availability

This study uses data from different sources. The whole slide images from the TCGA-BRCA study are publicly available at the GDC data portal: https://portal.gdc.cancer.gov/projects/TCGA-BRCA. The whole slide images from the Histo-AI, SmallTNBC, the segmentation annotations, the TSR scores of human experts in the TCGA-BRCA and the SmallTNBC datasets are subject to availability upon request. Please contact the corresponding author for access.

## Code Availability

We guarantee our code will be publicly available upon publication of this manuscript.

## Acknowledgements

We acknowledge Wangzhao Song for annotating the tissue subtypes in the TCGA-BRCA and the Histo-AI datasets used for training the segmentation model. The authors acknowledge all the staff of the core facility molecular pathology & biobanking (CFMPB) at the Netherlands Cancer Institute for making the slides from the Histo-AI available for this study. The authors acknowledge the staff at the research high performance computing (RHPC) facility for maintaining the compute clusters used in this study to train the AI models.

## Authorship

AP: Literature research, data collection, data curation, data interpretation, data analysis, model development, software development, study design, creation of figures and writing. JB: Supervision, writing, data interpretation, model development, software development and review. EM: Supervision, model development, software development and review. JS: Manual scoring of TSR in TCGA-BRCA dataset. MSG: Manual scoring of TSR in TCGA-BRCA and SmallTNBC. RM: Statistical analyses, data interpretation, writing and review. CIS: Supervision, project administration, study design and review. HMH: Supervision, project administration, funding acquisition, study design, review, manual scoring of TSR in TCGA-BRCA, data collection, data acquisition, data interpretation. JT: Supervision, project administration, funding acquisition, study design, software development, and review.

## References

1. Bremnes, R. M. et al. The role of tumor stroma in cancer progression and prognosis: emphasis on carcinoma-associated fibroblasts and non-small cell lung cancer. J. thoracic oncology 6, 209–217 (2011).

2. Panayiotou, H. et al. The prognostic significance of tumour-stroma ratio in endometrial carcinoma. BMC cancer 15, 1–8 (2015).

3. Chen, Y., Zhang, L., Liu, W. & Liu, X. Prognostic significance of the tumor-stroma ratio in epithelial ovarian cancer. BioMed research international 2015, 589301 (2015).

4. Scheer, R. et al. Tumor-stroma ratio as prognostic factor for survival in rectal adenocarcinoma: A retrospective cohort study. World journal gastrointestinal oncology 9, 466 (2017).

5. Lee, D. et al. Intratumor stromal proportion predicts aggressive phenotype of gastric signet ring cell carcinomas. Gastric Cancer 20, 591–601 (2017).

6. Vangangelt, K. et al. Prognostic value of tumor–stroma ratio combined with the immune status of tumors in invasive breast carcinoma. Breast cancer research treatment 168, 601–612 (2018).

7. Sullivan, L., Pacheco, R. R., Kmeid, M., Chen, A. & Lee, H. Tumor stroma ratio and its significance in locally advanced colorectal cancer. Curr. Oncol. 29, 3232–3241 (2022).

8. Karancsi, Z. et al. Tumour-stroma ratio (tsr) in breast cancer: comparison of scoring core biopsies versus resection specimens. Virchows Arch. 1–14 (2023).

9. de Kruijf, E. M. et al. Tumor–stroma ratio in the primary tumor is a prognostic factor in early breast cancer patients, especially in triple-negative carcinoma patients. Breast cancer research treatment 125, 687–696 (2011).

10. Micke, P. et al. The prognostic impact of the tumour stroma fraction: A machine learning-based analysis in 16 human solid tumour types. EBioMedicine 65 (2021).

11. Zheng, Q. et al. Machine learning quantified tumor-stroma ratio is an independent prognosticator in muscle-invasive bladder cancer. Int. J. Mol. Sci. 24, 2746 (2023).

12. Hong, Y. et al. Deep learning-based virtual cytokeratin staining of gastric carcinomas to measure tumor–stroma ratio. Sci. Reports 11, 19255 (2021).

13. Smit, M. A. et al. Deep learning based tumor–stroma ratio scoring in colon cancer correlates with microscopic assessment. J. Pathol. Informatics 14, 100191 (2023).

14. Zhao, K. et al. Artificial intelligence quantified tumour-stroma ratio is an independent predictor for overall survival in resectable colorectal cancer. EBioMedicine 61 (2020).

15. Beck, A. H. et al. Systematic analysis of breast cancer morphology uncovers stromal features associated with survival. Sci. translational medicine 3, 108ra113–108ra113 (2011).

16. Mesker, W. E. et al. The carcinoma–stromal ratio of colon carcinoma is an independent factor for survival compared to lymph node status and tumor stage. Anal. Cell. Pathol. 29, 387–398 (2007).

17. van Pelt, G. W. et al. Scoring the tumor-stroma ratio in colon cancer: procedure and recommendations. Virchows Arch. 473, 405–412 (2018).

18. Hagenaars, S. C. et al. Standardization of the tumor-stroma ratio scoring method for breast cancer research. Breast cancer research treatment 193, 545–553 (2022).

19. Amgad, M. et al. Report on computational assessment of tumor infiltrating lymphocytes from the international immuno-oncology biomarker working group. NPJ breast cancer 6, 16 (2020).

20. Sikaroudi, M., Hosseini, M., Gonzalez, R., Rahnamayan, S. & Tizhoosh, H. Generalization of vision pre-trained models for histopathology. Sci. reports 13, 6065 (2023).

21. Howard, F. M. et al. The impact of site-specific digital histology signatures on deep learning model accuracy and bias. Nat. communications 12, 4423 (2021).

22. Howard, F. M., Kather, J. N. & Pearson, A. T. Multimodal deep learning: an improvement in prognostication or a reflection of batch effect? Cancer Cell 41, 5–6 (2023).

23. Lennerz, J. K., Green, U., Williamson, D. F. & Mahmood, F. A unifying force for the realization of medical ai. NPJ Digit. Medicine 5, 172 (2022).

24. Klauschen, F. et al. Scoring of tumor-infiltrating lymphocytes: From visual estimation to machine learning. In Seminars in cancer biology, vol. 52, 151–157 (Elsevier, 2018).

25. Schirris, Y. et al. Weakstil: Weak whole-slide image level stromal tumor infiltrating lymphocyte scores are all you need. In Medical Imaging 2022: Digital and Computational Pathology, vol. 12039, 55–59 (SPIE, 2022).

26. Wetstein, S. C. et al. Deep learning-based breast cancer grading and survival analysis on whole-slide histopathology images. Sci. reports 12, 15102 (2022).

27. Jaroensri, R. et al. Deep learning models for histologic grading of breast cancer and association with disease prognosis. NPJ Breast cancer 8, 113 (2022).

28. Mercan, C. et al. Deep learning for fully-automated nuclear pleomorphism scoring in breast cancer. NPJ breast cancer 8, 120 (2022).

29. Shamai, G. et al. Deep learning-based image analysis predicts pd-l1 status from h&e-stained histopathology images in breast cancer. Nat. Commun. 13, 6753 (2022).

30. Polley, M.-Y. C. et al. An international study to increase concordance in ki67 scoring. Mod. pathology 28, 778–786 (2015).

31. Nielsen, K. et al. High inter-laboratory variability in the assessment of her2-low breast cancer: a national registry study on 50,714 danish patients. Breast Cancer Res. 25, 139 (2023).

32. Kofler, F. et al. Are we using appropriate segmentation metrics? identifying correlates of human expert perception for cnn training beyond rolling the dice coefficient. Mach. Learn. for Biomed. Imaging 2, 27–71 (2023).

33. Reinke, A. et al. Understanding metric-related pitfalls in image analysis validation. Nat. methods 21, 182–194 (2024).

34. Lovchinsky, I. et al. Discrepancy ratio: evaluating model performance when even experts disagree on the truth. In International Conference on Learning Representations (2019).

35. Oktay, O. et al. Attention u-net: Learning where to look for the pancreas. arXiv preprint arXiv:1804.03999 (2018).

36. Liu, J. et al. An integrated tcga pan-cancer clinical data resource to drive high-quality survival outcome analytics. Cell 173, 400–416 (2018).

37. Amgad, M. et al. Structured crowdsourcing enables convolutional segmentation of histology images. Bioinformatics 35, 3461–3467 (2019).

38. SS, B. Slide score. less scoring, more research! https://www.slidescore.com/. [Accessed: 17-Jul-2024].

39. Vallat, R. Pingouin: statistics in python. J. Open Source Softw. 3, 1026, DOI: 10.21105/joss.01026 (2018).

40. Koo, T. K. & Li, M. Y. A guideline of selecting and reporting intraclass correlation coefficients for reliability research. J. chiropractic medicine 15, 155–163 (2016).

