## Supplementary Material for "A quantitative comparison between human experts and AI at estimating tumor-stroma ratio"

### 1 Model training details

##### Training and validation data

For training, we used data from 156 patients, totaling 166 WSIs (144 Aperio images and 22 P1000 images). The validation set consisted of data from 76 patients with 79 WSIs (72 Aperio and 7 P1000 images).

#### 1.1 Data annotation

On the aforementioned WSIs utilised in this work, we obtained detailed ground truth annotations for various tissue types. They include: stroma, tumor, inflamed regions, ductal carcinoma in-situ (DCIS) regions, regions with lymphoid aggregate, regions with DCIS immune cells, necrotic tissue, normal glands, regions with blood vessels, fat cell regions, regions with red blood cells, fibrotic tissue, artefacts (such as tissue folding, tears, out of focus etc), background area, skin tissue, liver tissue and unlabelled area.

Of these, we remap tissue-types into three distinct classes. The first class is stroma containing stroma, inflamed regions, regions with lymphoid aggregate, regions with DCIS immune cells, normal glands, regions with blood vessels, regions with red blood cells, fibrotic tissue. The second class is tumor containing tumor and DCIS regions. The final class is of all other components including necrotic tissue, fat cell regions, background area, artefacts, skin tissue, liver tissue, and unlabelled area. prior guidelines were followed while remapping the tissue types[1].

#### 1.2 Tissue segmentation model

We use the open source digital pathology deep-learning framework 'ahcore' developed at the Netherlands Cancer Institute. Within this, we employed an Attention U-Net model[2] from the MONAI library[3]. The architecture includes five encoding layers, configured with feature channels increasing per layer as [32, 64, 128, 256, 512, 1024]. Each layer includes a stride of 2 for down-sampling, and a dropout rate of 0.1 is applied to support regularization. Best model selection occurred based on the highest stroma Dice-Sørensen coefficient on the validation data. We used the standard cross-entropy loss as an optimization target, where non-annotated regions were ignored. Model training was conducted on a single NVIDIA A100 GPU with training batch size set to 32.

#### 1.3 Data pre-processing

We used the Deep Learning Utilities for Pathology (DLUP) library developed at the Netherlands Cancer Institute to tile WSIs into fixed-size samples suitable for deep learning. During training, the model accessed only annotated regions within these tiles, focusing learning on areas relevant to the task and optimizing memory use. Each batch yielded by the dataset was normalized to have zero mean and a standard deviation of one.

#### 1.4 Augmentations

We apply an extensive augmentation strategy, implemented through the Kornia library[4]. This strategy included both intensity-based and geometric transformations. For intensity augmentations, we utilized several techniques following prior work[5]: Gaussian blur with a probability of 0.5, kernel size of 9 by 9, and sigma range 0.1 to 1.0, sharpness with a probability of 0.5 and a sharpness factor of 10; and color jitter, also with a probability of 0.5, configured to adjust brightness (0.65 to 1.35), contrast (0.5 to 1.5), hue (-0.1 to 0.1), and saturation (0.5 to 0.9).

The geometric transformations were applied in arbitrary order to introduce spatial variability. These included horizontal flip and vertical flip, each with a probability of 0.5, as well as perspective distortion with a 0.5 probability and a distortion scale of 0.1. Additionally, several affine transformations were used: random rotations up to 90 degrees (p = 0.5), scaling within a range of 0.75 to 1.0 (p = 1.0), translations up to 5 percent of the image size (p = 0.5), and shear adjustments within a range of 0 to 5 degrees (p = 0.5).

For standardization, a final center crop was applied to each image, resizing it to 1024 by 1024 pixels. All these augmentations were applied exclusively during the training phase to increase the model's robustness to variations in the data. During validation, all intensity, geometric, and post-processing augmentations were disabled to ensure consistent evaluation conditions.

### 2 Determining the TSR ambiguity bounds using the Dice metric for a deep learning model

Our AI-based approach for TSR estimation produces point predictions, since the estimate is defined as a ratio of segmented pixel counts. However, segmentation models are generally imperfect, meaning that the generated pixel masks likely contain a set of wrongly classified pixels. This is captured by the Dice-Sørensen coefficient (DSC), defined in terms of the ground truth mask $T$ and the predicted mask $P$ as

$$D = \frac{2|T \cap P|}{|T \cap P| + |T \cup P|}. \quad (1)$$

Only a pixel-perfect segmentation mask leads to a DSC equal to 1, whereas a DSC of 0 implies that none of the ground-truth foreground pixels were correctly classified as foreground by the model (with an arbitrary number of false positives). If a segmentation model typically outputs masks with imperfect DSC metrics on validation data, we can therefore expect such a model to misclassify a certain number of pixels on unseen inputs in general. These misclassified pixels can affect the TSR score derived from the mask, meaning that it is inappropriate to view these scores as true point predictions. To rephrase: acknowledging that our segmentation model is imperfect forces us to accept that there is some ambiguity in the AI-derived TSR prediction.

We can use this observation to derive upper and lower bounds on the AI-derived TSR estimate that are still consistent with a given DSC value. This leads to a quantitative, but *heuristic* notion of 'uncertainty' on the AI-derived TSR estimate, which we refer to as the *TSR ambiguity bounds*. We emphasize that these bounds do not correspond to any standard *statistical* notions of confidence intervals or error bars, and should not be interpreted as such. They are merely a rough order-of-magnitude estimate of the ambiguity involved in the predicted TSR value, taking into account a certain degree of imperfection in the segmentation performance.

In order to derive this estimate, we make the simplifying assumption that the model attains a certain fixed DSC value on all unseen inputs. This fixed DSC value could, for example, be chosen as the mean of the DSC (for a given tissue compartment) over the entire validation set (with available ground truth segmentation masks) for each tissue compartment type (e.g. tumor or stroma). Clearly, this assumption is typically violated to a certain degree in practice, but we can ignore this for the purpose of deriving our order-of-magnitude estimate of the bounds.

We can then ask what maximally oversegmented or maximally undersegmented masks are still consistent with this fixed DSC value. By 'maximum oversegmentation' we mean that all misclassified pixels are false positives (and zero false negatives), whereas by 'maximum undersegmentation' we mean that all misclassified pixels are false negatives (and zero false positives). We refer to Figure 1 for a diagrammatic representation of these two scenarios. For a given tissue compartment, the maximum oversegmentation case therefore leads to the maximal number of predicted 'foreground' pixels that is still consistent with the associated prespecified DSC value, and the minimal number of such pixels in the case of maximal undersegmentation. This leads to maximum and minimum pixel counts for each tissue compartment.

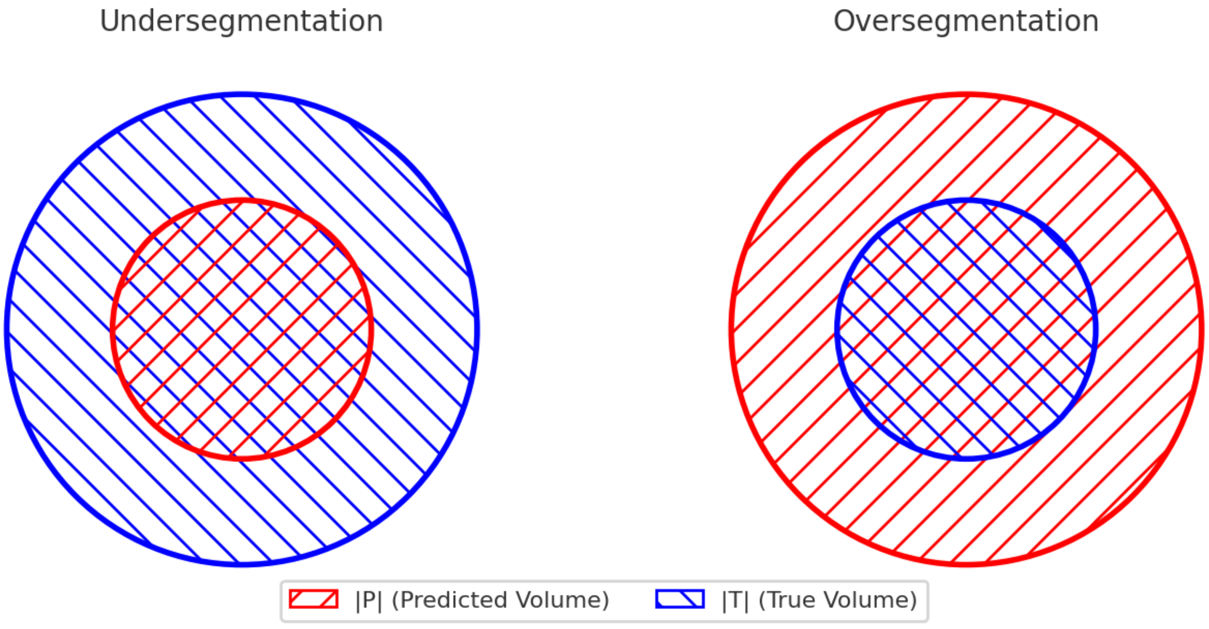


**Figure 1.** Illustration of maximum undersegmentation and maximum oversegmentation.

Since the TSR is defined as a ratio of such pixel counts, we use the respective maximum and minimum pixel counts to derive our heuristic TSR bounds. From Equation 1, we see that the upper bound for TSR occurs in the case of maximal oversegmentation of stroma and maximal undersegmentation of tumor, whereas the lower bound occurs in the case of maximal undersegmentation of stroma and maximal oversegmentation of tumor.

Before we proceed to derive the expressions for the TSR bounds, we first define the pixel count variables $I = |T \cap P|$ and $U = |T \cup P|$ (note that these are the pixel counts of the intersection and union, respectively, of the ground truth mask $T$ and the predicted mask $P$). Furthermore, from de Morgan's laws we have that $U = |T| + |P| - |T \cap P|$.

Next, we express the size of the maximal over- and undersegmentation in terms of the DSC and the ground truth mask size $|T|$.

##### 2.1 Oversegmentation

Since all segmentation errors are false positives, we can write $|P| = |T| + \varepsilon_{\text{over}}$, where $\varepsilon_{\text{over}}$ is the number of these false-positive pixels. Furthermore, we have that $I = |T| = |P| - \varepsilon_{\text{over}}$ and $U = |P|$. From (1) and the aforementioned identities, we find

$$\begin{aligned}\frac{2}{D} &= \frac{I+U}{I} \\ &= \frac{2|P| - \varepsilon_{\text{over}}}{|P| - \varepsilon_{\text{over}}},\end{aligned}$$

so that

$$\varepsilon_{\text{over}} = \frac{2|P|(1-D)}{2-D}.$$

##### 2.2 Undersegmentation

Now we have that $|P| = |T| - \varepsilon_{\text{under}}$, $I = |P|$, and $U = |P| - \varepsilon_{\text{under}}$. We find

$$\frac{2}{D} = \frac{2|P| + \varepsilon_{\text{under}}}{|P|}$$

and therefore

$$\varepsilon_{\text{under}} = \frac{2|P|(1-D)}{D}.$$

##### 2.3 TSR ambiguity bounds

Let $V_S$ and $V_T$ denote the stroma and tumor volumes, respectively, as segmented by the AI model within the most invaded region (MIR) visible on a whole slide image (WSI) of a breast tumor resection. Since $V_S$ and $V_T$ are likely based on imperfect pixel classifications, the resulting volumetric measurements remain ambiguous. To bracket the ambiguity in quantifying $V_S$ and $V_T$, we compute maximal error margins as noted above, denoted by $\varepsilon_{\text{over},C}$, $\varepsilon_{\text{under},C}$ from the validation DSC value $D_C$, where $C$ is either $S$ or $T$.

Finally, we can proceed to define the upper and lower bounds for the AI derived TSR estimates consistent with validation Dice scores $D_{S,T}$. Given these values, the TSR upper bound is obtained by adding the maximum stroma oversegmentation error ($\varepsilon_{\text{over},S}$) to $V_S$ and subtracting the maximum tumor undersegmentation error ($\varepsilon_{\text{under},T}$) from $V_T$ in Equation 1:

$$\text{TSR}_{\text{upper}} = \frac{V_S + \varepsilon_{\text{over},S}}{(V_S + \varepsilon_{\text{over},S}) + (V_T - \varepsilon_{\text{under},T})}.$$

Similarly, to obtain the TSR lower bound, we subtract the maximum stroma undersegmentation error ($\varepsilon_{\text{under, S}}$) from $V_S$ and add the maximum tumor oversegmentation error to $V_T$ in Equation 1:

$$\text{TSR}_{\text{lower}} = \frac{V_S - \varepsilon_{\text{under},S}}{(V_S - \varepsilon_{\text{under},S}) + (V_T + \varepsilon_{\text{over},T})}.$$

In terms of the fixed DSC values ($D_S$ for stroma and $D_T$ for tumor), we obtain

$$\text{TSR}_{\text{upper}} = \frac{V_S\left(\frac{4-3D_S}{2-D_S}\right)}{V_S\left(\frac{4-3D_S}{2-D_S}\right) + V_T\left(\frac{3D_T-2}{V_T}\right)}, \tag{2}$$

$$\text{TSR}_{\text{lower}} = \frac{V_S\left(\frac{3D_S-2}{D_S}\right)}{V_S\left(\frac{3D_S-2}{D_S}\right) + V_T\left(\frac{4-3D_T}{2-D_T}\right)}. \tag{3}$$

#### 3 Limitations of human and AI estimates of TSR

First, we present cases that show the limitations of human evaluation in accurately quantifying TSR when compared to the DL model. Figure 2 demonstrates an example from the SmallTNBC dataset where the visual contrast between the stromal and tumor compartments is inherently poor. In this instance, the stromal compartment interspersed with the tumor compartment is stained in a manner visually similar to tumor cells. For the human observer, accurately distinguishing these regions is harder since it requires meticulous attention to the differences in morphology. In contrast, the AI model identifies textural and structural features to effectively distinguish between these compartments, providing a more reliable segmentation.

Similarly, Figure 3 shows a case from the TCGA-BRCA dataset where human estimation of TSR is compromised due to ambiguity in differentiating the stromal compartment. Factors such as overlapping cellular structures, irregular compartmentalization make it challenging to accurately assess the stromal regions. On the other hand, the complicated stromal region is reliably assessed by the AI model.

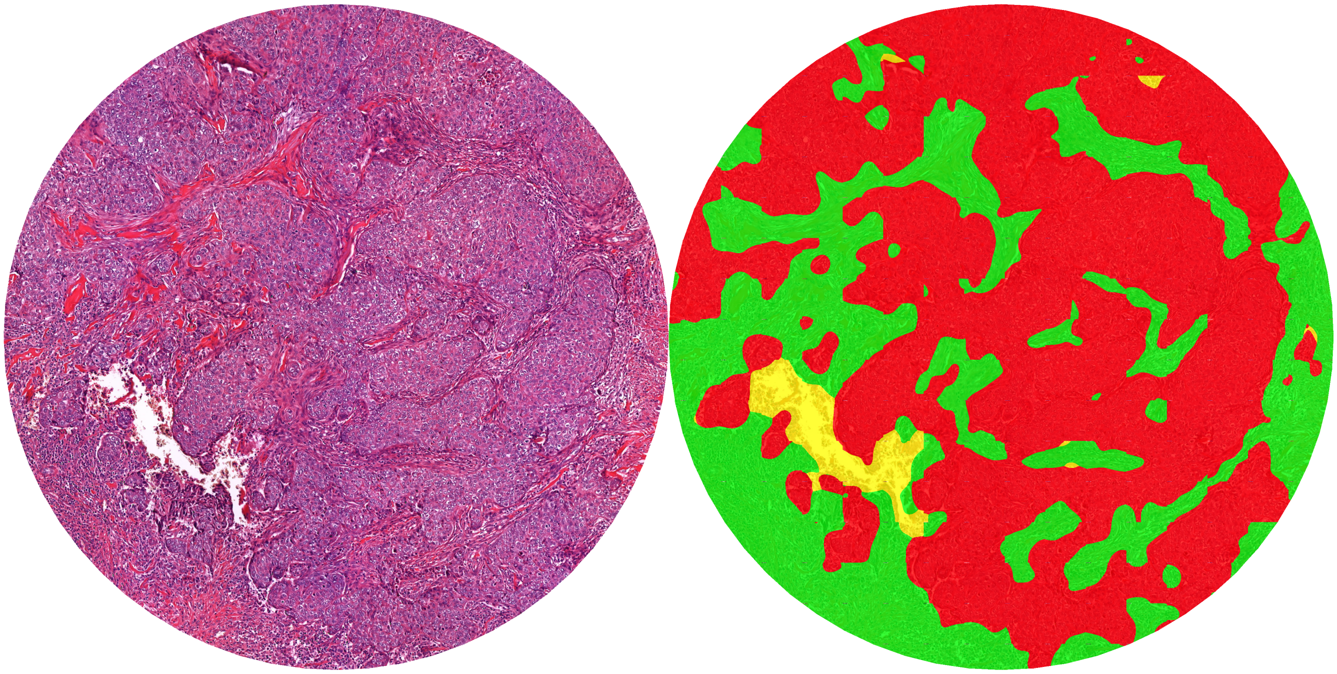


**Figure 2.** Failure in human estimation of TSR due to poor contrast in staining. When the AI-segmentation is shown side-by-side, the human error becomes apparent.

Second, we present examples that highlight the limitations of the AI model in quantifying TSR, illustrating scenarios where its performance is compromised. Figure 4 illustrates a scenario where the model underperforms due to a tumor morphology rarely seen in the training dataset. The limited representation of such rare morphologies in the training data hampers the model's ability to generalize to these cases, resulting in segmentation errors and consequently in inaccurate TSR estimation.

Figure 5 depicts an example where the AI model overestimates the tumor area. In this case, the fine-grained details of the stroma are missed, leading to an oversegmentation of the tumor compartment. This misclassification reduces the precision of TSR estimation and highlights the model's difficulty in capturing nuanced tissue boundaries when the contrast between compartments is subtle. On the other hand, the AI model correctly identifies the blurring artefact in the bottom left corner, which is easily missed by human eyes.

Figure 6 shows another challenging case where the AI model fails to recognize tissue tears, misclassifying them as tumor. The disruption introduced by the tears creates a visual pattern that the model has not learned to interpret correctly, resulting in an inaccurate assignment of tissue labels. Such errors underscore the importance of robust training on a wider range of artifact and tissue variations to improve the reliability of AI-based TSR quantification.

#### References


1. Hagenaars, S. C. *et al.* Standardization of the tumor-stroma ratio scoring method for breast cancer research. *Breast cancer research treatment* **193**, 545–553 (2022).

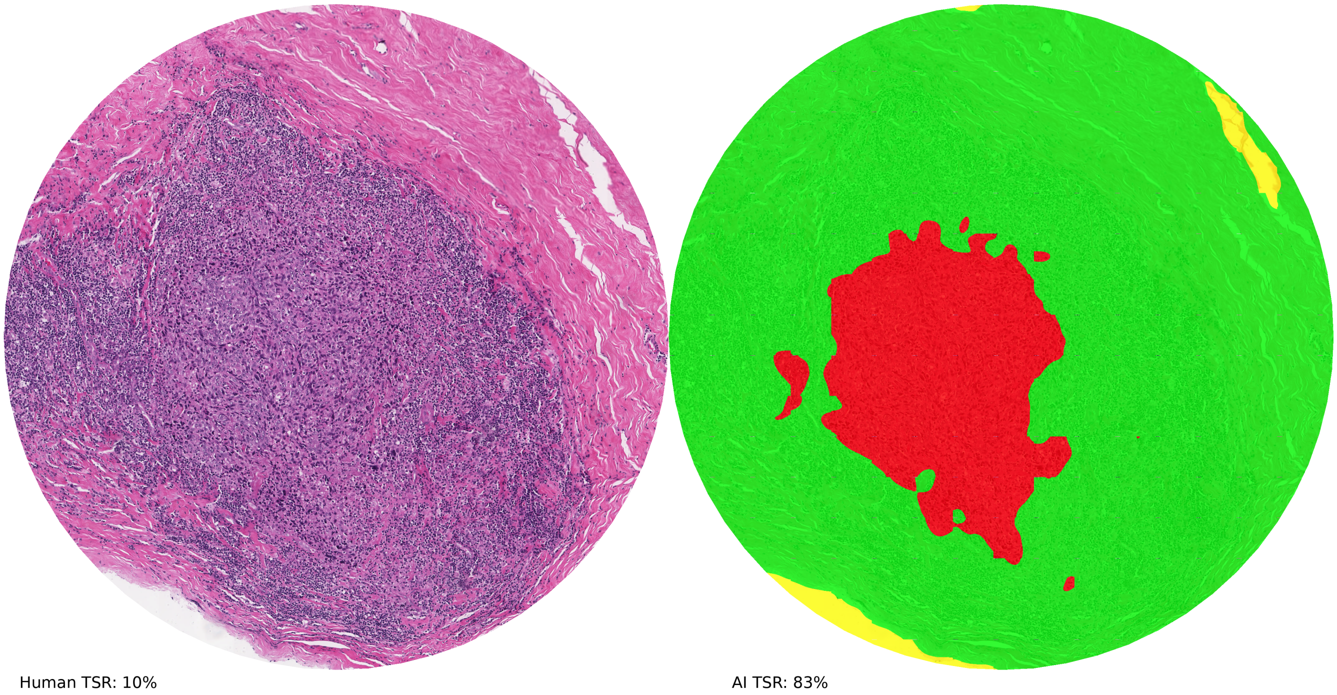


**Figure 3.** Failure in human estimation of TSR due to difficulty in distiguishing between stroma and tumor. In this example, lymphocytes are confused for tumor cells by the human.


**2.** Oktay, O. *et al.* Attention u-net: Learning where to look for the pancreas. *arXiv preprint arXiv:1804.03999* (2018).

**3.** Cardoso, M. J. *et al.* Monai: An open-source framework for deep learning in healthcare (2022). 2211.02701.

**4.** Riba, E., Mishkin, D., Ponsa, D., Rublee, E. & Bradski, G. Kornia: an open source differentiable computer vision library for pytorch. In *Winter Conference on Applications of Computer Vision* (2020).

**5.** Tellez, D. *et al.* H and e stain augmentation improves generalization of convolutional networks for histopathological mitosis detection. In *Medical Imaging 2018: Digital Pathology*, vol. 10581, 264–270 (SPIE, 2018).

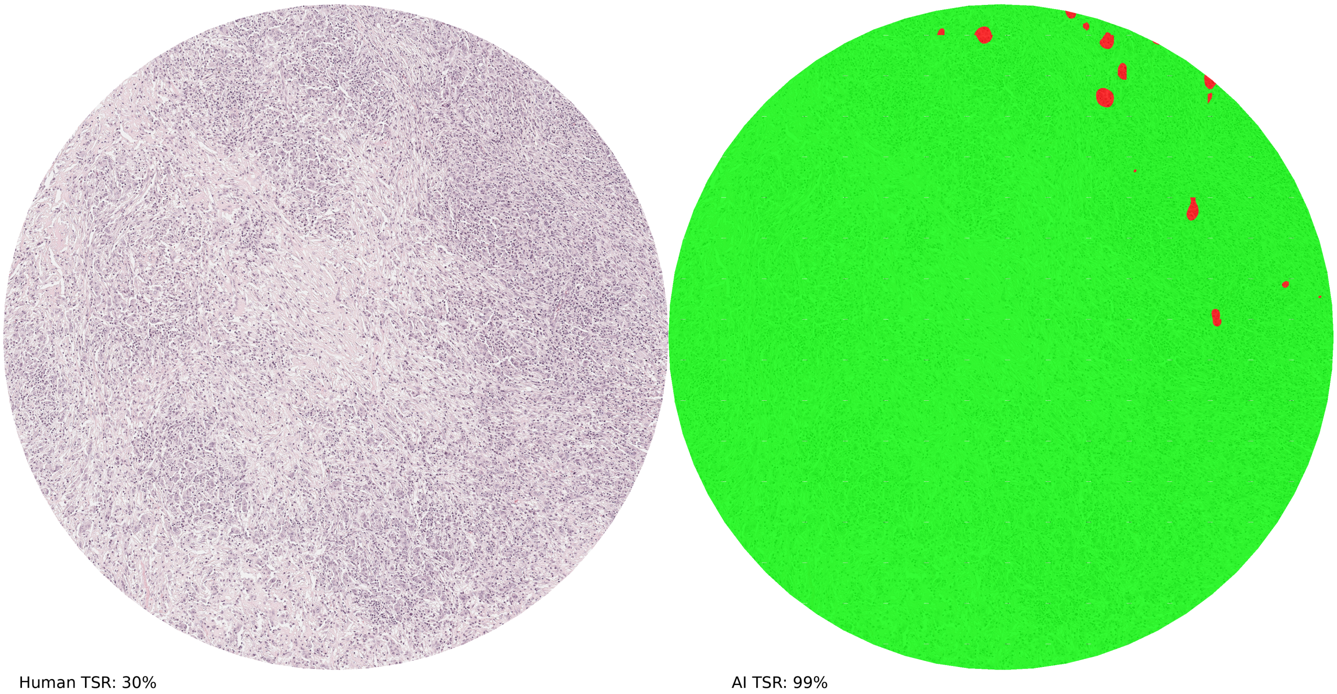


**Figure 4.** Failure of AI model to capture tumor morphology alterations.

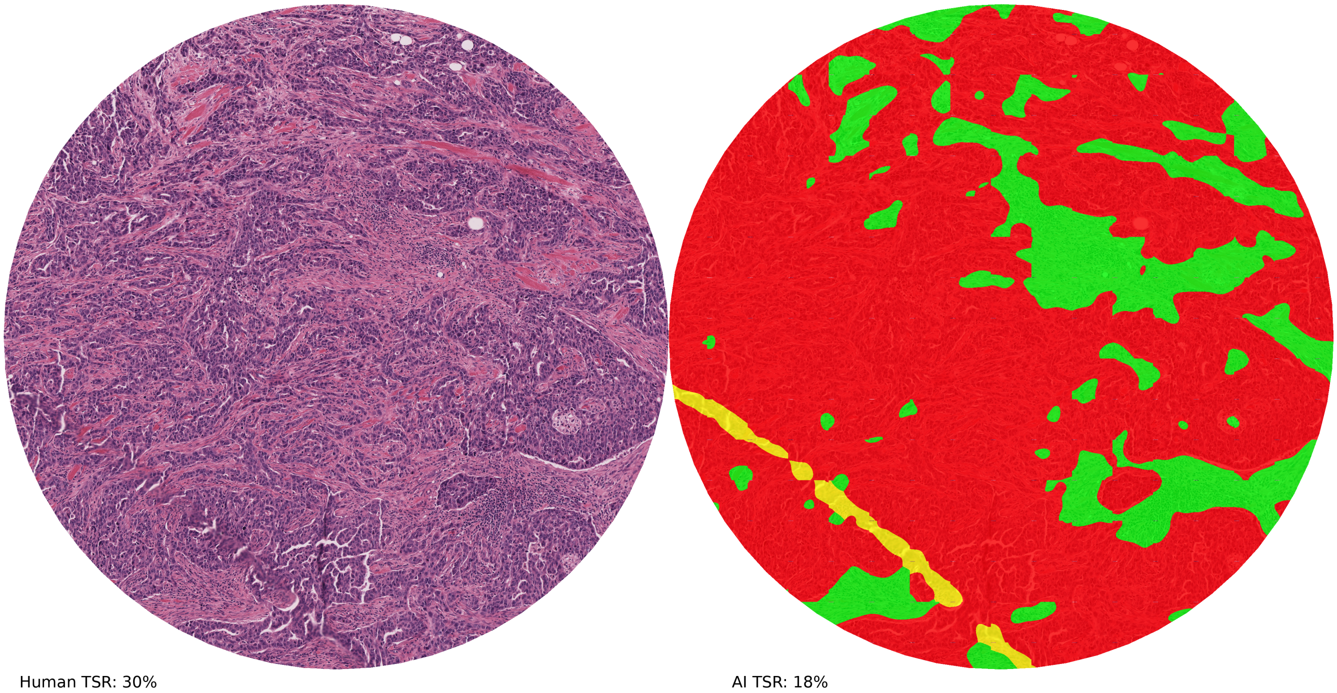


**Figure 5.** Overestimation of tumor by the AI model. Fine grained details of stroma are missed by the AI model during segmentation. The blurring artefacts are correctly identified by the AI model.

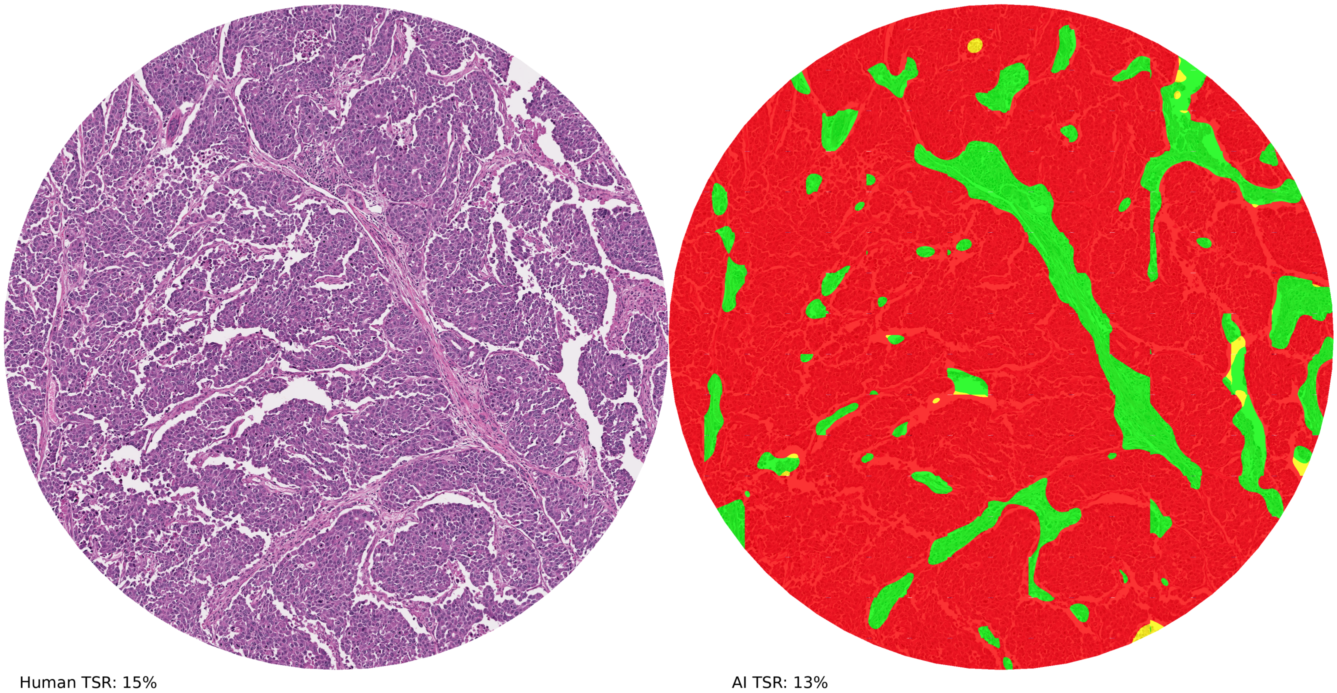


**Figure 6.** Failure of AI model to capture tissue tears. The model classifies tears as tumor in this example.
